# An ecological study of the effect of white-tailed deer on alpha-gal syndrome in United States counties

**DOI:** 10.64898/2026.08.13.26360354

**Authors:** Marco Piccininni, Luis Cadahía, Mats J. Stensrud

## Abstract

**Background:** Alpha-gal syndrome (AGS) is an emerging disease, increasingly recognized as a public health concern in the United States. The primary cause of AGS in the United States is the bite of *Amblyomma americanum* ticks. White-tailed deer serve both as a preferred food source and as transport for *A. americanum*. In this work, we aim to quantify the effect of white-tailed deer abundance on number of AGS cases in United States counties.

**Methods:** To mitigate concerns about confounding, we used the front-door formula, leveraging biological knowledge about the causal process. Due to lack of official data, our analysis relied on data made available by citizen science efforts.

**Results:** We found that a higher number of reported white-tailed deer sightings in 2020 was associated with the county-level presence of *A. americanum* in 2024. In turn, county-level presence of *A. americanum* was associated with a higher number of self-reported AGS cases. We estimated that if white-tailed deer abundance had increased by 50%, 75%, or 100% in 2020, there would have been 89 (95%CI: 12, 252), 126 (12, 354) or 159 (6, 448) additional AGS self-reported cases in the US in 2025.

**Conclusions:** The estimated associations are compatible with an effect of white-tailed deer abundance on AGS in the country. Due to measurement error, the low granularity of the available data, the ecological nature of the design, and the modelling choices, our effect estimates should be interpreted cautiously. Further studies are needed to quantify the population-level effect of white-tailed deer on AGS.

**Impact statement:** In this ecological study of United States counties, the authors found associations compatible with an effect of white-tailed deer abundance and alpha-gal syndrome prevalence.

## Introduction

Alpha-gal syndrome (AGS), also known as “red meat” allergy^1,2^, is an emerging disease characterized by potentially severe allergic symptoms that appear after consuming mammalian meat or mammalian-derived products^1–10^. The name alpha-gal is short for galactose-alpha-1,3-galactose, an oligosaccharide present in most mammals, but absent in humans and some other primates^1,2,5,9,11^. The description of AGS emerged from investigations into unusual allergic reactions in the early 2000s^12–14^, attributed to the presence of IgE antibodies against alpha-gal^1,2,5–7,11^.

It has been suggested that tick bites lead to alpha-gal specific IgE sensitization, which can later cause AGS^1–3,5–8,10,11^. Indeed, alpha-gal is an endogenous tick component that has been found in the salivary glands, saliva, gastrointestinal track and hemolymph^2,7,11^.

In the US, most cases are reported in the southern, midwestern, and mid-Atlantic regions^2,15^. The primary cause for the increase in alpha-gal specific IgE sensitization in these areas is believed to be bites of *Amblyomma americanum* ticks, also referred to as lone star ticks^1–3,5–7,10,11,15^. The sensitization by the *A. americanum* tick is supported by case reports, ecological studies, and observational studies^1,5,6,8,11,15^. Further evidence comes from experimental studies in mice^9,16^.

*A. americanum* is a widely distributed, hard-bodied tick whose keystone wildlife host is the American white-tailed deer (*Odocoileus virginianus*), which supports all parasitic stages of this tick species^17^. White-tailed deer serve both as a preferred food source and as transport^17^. In favorable habitats, they can support very high numbers of *A. americanum* ticks^17,18^. The association between abundance of white-tailed deer and *A. americanum* is supported by mathematical models^17,19^ and by deer exclusion studies^17^. White-tailed deer populations have expanded substantially in the eastern US during the twentieth century^11,17^. This is likely to have brought about an expansion of the range of *A. americanum* ticks^17^ and, with it, a higher risk of tick exposure and development of AGS^4,11,20^. Recent studies show an increase in the number of AGS cases between 2010-2018^8^ and 2017-2022^15^.

Despite the connection between the white-tailed deer and the *A. americanum* tick, the effect of the increasing deer abundance on the development of AGS has received little attention. In this work, we aimed to quantify the effect of white-tailed deer abundance on the number of AGS cases in US counties. Such causal questions are difficult to answer because of lack of official and granular data, and the incomplete understanding of AGS development.

## Methods

### Motivation for the analytical strategy

Causal questions about the effect of white-tailed deer on AGS are not easy to answer. Ideally, such questions should be studied by running randomized trials where individuals living in the US are randomly assigned to live in identical areas with different abundances of white-tailed deer. Such experiments are obviously infeasible. Alternatively, we could attempt to answer such questions with observational cohort data containing individual level measurements of residential location, deer exposure, incidence of AGS, and relevant confounder variables. Unfortunately, this type of individual-level data is scarce and difficult to obtain. Rather than consider individual-level data, we therefore conducted an ecological study. We studied the effect of white-tailed deer abundance on AGS at the US county level, conceptualizing counties as random draws from a very large superpopulation.

Another major difficulty in answering our research question is that relatively little is known about AGS, so comparisons between counties are prone to confounding. Experts do not yet understand the precise pathological mechanisms of AGS and what the possible causes of this disease are^1,2,4,10^. Therefore, comparing AGS cases between counties that have a low abundance of white-tailed deer with counties that have a higher abundance can be misleading. Besides obvious confounders, such as the size of the county and the number of humans living in the region, the counties with higher abundance of white-tailed deer are in regions with particular socio-economic factors, history, traditions, and environmental characteristics. Thus, a naive comparison could misleadingly attribute differences in AGS occurrence to white-tailed deer abundance, due to other factors (e.g., different dietary patterns, different use of pharmaceutical drugs, environmental differences).

To minimize concerns about unmeasured confounding of the relationship between white-tailed deer abundance and AGS, we use the front-door strategy, grounded on formal causal inference results^21–23^. This analytic strategy allows for the identification of the causal effect of an exposure on an outcome, even when exposure-outcome unmeasured confounding exists, if information from a variable that fully mediates the causal effect is available^21–24^. The basic idea behind the front-door strategy is that we can first estimate the effect of the exposure on what we call the full mediator, a variable that entirely mediates the exposure’s effect. Then we estimate the effect of the full mediator on the outcome, and finally combine these two effects to obtain the effect of the exposure on the outcome^22,23^. The front-door formula is rarely used in health research^25,26^, but has been praised by methodologists for giving valid results when standard methods fail^23^.

In our study, the front-door formula is particularly suitable because we know that the main mechanism, if not the only mechanism, through which deer abundance affects AGS occurrence is the level of *A. americanum* presence in the county^11,20^. Recently two case reports suggested that other tick species, such as *Ixodes scapularis*^27^ and *Ixodes pacificus*^28^, which can be transported by white-tailed deer as well, could transmit AGS in the US. However, the genus *Ixodes* has yet to be conclusively linked with AGS in the US^28^, and the role of these other ticks seems negligible compared to the role of *A. americanum*^28^. Therefore, we consider the presence of *A. americanum* in the county as the full mediator variable.

Variables such as the size of the human population, area of the county, temperature, and total precipitation can affect deer abundance, *A. americanum* presence, and the number of AGS cases. For this reason, we also consider these variables in the analysis. In the causal graph^21–23^ in Figure 1 we represent the idealized causal model. Here, we focus on the effect of white-tailed deer abundance in 2020 on AGS cases in 2025.

**Figure 1.**
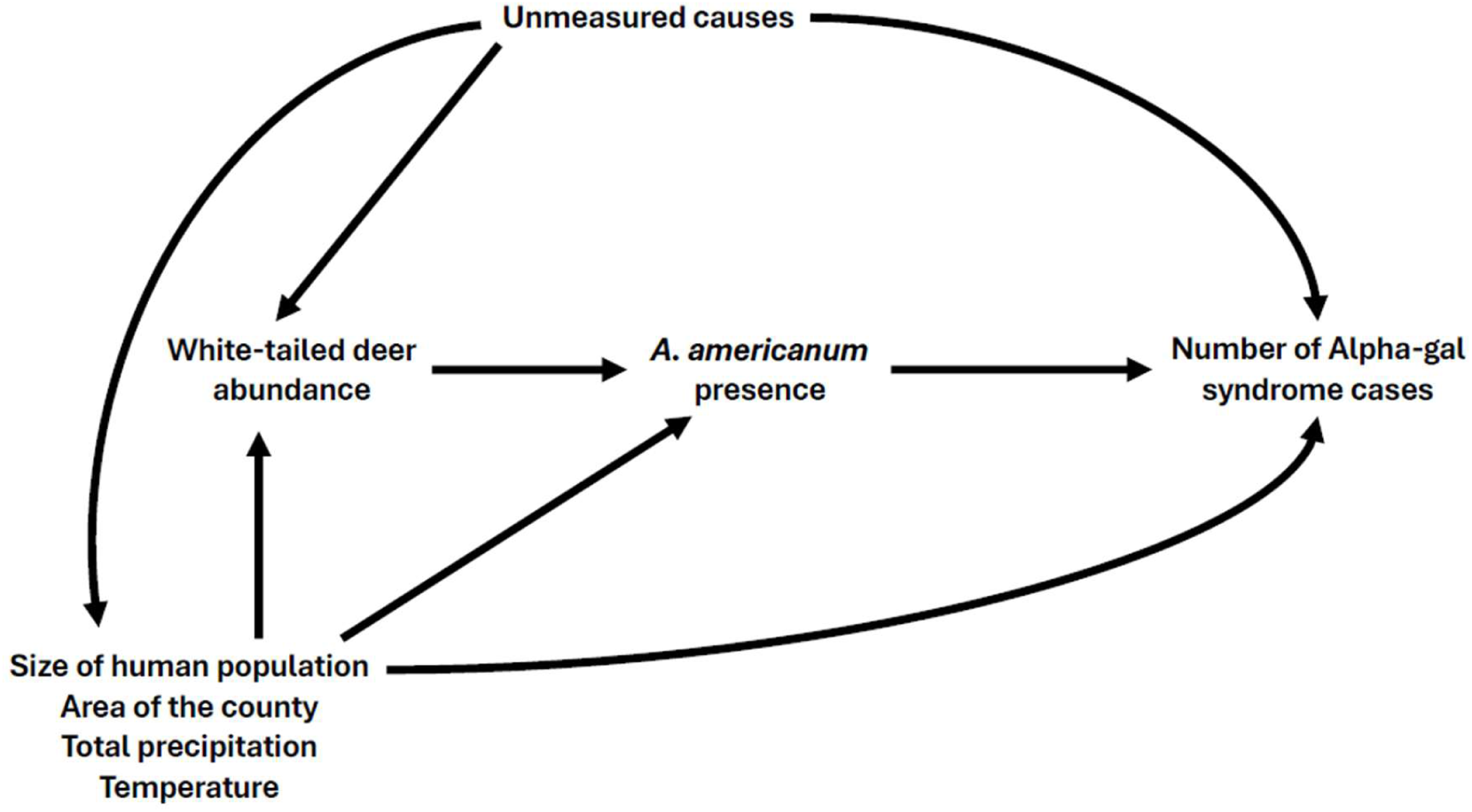
Causal directed acyclic graph representing the idealized data generating process.

### Data

We considered US counties and county equivalents in the conterminous United States. Estimates for the number of persons living in each US county in 2024 according to the U.S. Census Bureau were downloaded from StatsAmerica’ s Download Center^29^. Because human population size does not change substantially over a five year period, we used this value for our analysis. TIGER/Line shapefiles and information about the total area of each county from the U.S. Census Bureau were obtained using the “tigris” R package^30^. Data about temperature and precipitation for each county were obtained from the National Oceanic and Atmospheric Administration using the “EpiNOAA” R package^31^. Highest and lowest temperatures, as well as amount of precipitation, are important predictors of *A. americanum* presence^32^. Thus, in our analysis we considered the maximum and minimum temperatures throughout 2020 and the total precipitation in 2020.

No official data on the abundance of *A. americanum* by US county is available. Therefore, we decided to use county-level surveillance evidence of presence as a proxy. The estimated presence of *A. americanum* through 2024 in each US county was downloaded from the Lone Star Tick Surveillance website of the Centers for Disease Control and Prevention (CDC)^33^.

Using a variety of sources, the CDC surveillance program reports three levels of *A. americanum* presence: No records, Reported, and Established^33^. A county is labelled as “Established” if six or more ticks of a single life stage or more than one life stage were collected in the county within a 12-month period, in the current or previous years^33^. Minor inconsistencies between datasets, such as those resulting from changes to county definitions in Connecticut^34^, were manually resolved.

Data about abundance of white-tailed deer and the number of AGS cases by county are also difficult to obtain. The exact prevalence of alpha-gal syndrome by county is unknown^8,15^. As a proxy for these quantities, we relied on open data made available by citizen science efforts.

As a proxy for white-tailed deer abundance, we used data on the number of reported human observations of white-tailed deer. Specifically, we used data from the Global Biodiversity Information Facility (GBIF)^35^. We extracted data on all human observations of *Odocoileus virginianus (Zimmermann, 1780)* (commonly known as white-tailed deer) in the United States in 2020 registered through the iNaturalist app^36^. iNaturalist is a widely used citizen science online social network that also functions as an organism occurrence recording tool^37^.

Sightings of white-tailed deer were attributed based on their latitude and longitude to the closest match US county. We considered the total number of reported sightings (irrespective of the number of animals) throughout 2020 in the county.

As a proxy for the number of AGS cases for each US county, we used self-reported data. As part of their Alpha-gal Syndrome Awareness Campaign, the Alpha-gal Alliance developed an Alpha-gal Information website^38^ where users with AGS around the world are invited to report their location on a map^39^. While we are unaware of the identity of the creator of the map, this crowdsourced map collected for years information about self-reported location of individuals with AGS. Thousands of pins have been added to the map in the US^40^, providing information about the geographic distribution of this understudied disease. Individuals with AGS who self-reported being in the US were then attributed to the closest county based on the latitude and longitude of their pin. Individuals who did not specify the country were attributed to a US county if their pin was within the county borders.

Data about temperature and precipitation were downloaded on June 20th, 2026. All other datasets were downloaded on December 31st, 2025.

### Statistical analysis

Our statistical analysis relies on the use of proxy variables. Specifically, we consider the cumulative number of self-reported AGS cases at the end of 2025 as outcome variable. We assume that the presence of *A. americanum* through 2024, as estimated by the CDC, is sufficient to fully capture the effect of 2020 white-tailed deer abundance, so that the causal model in Figure 1 holds true. We also assume that, after accounting for deer abundance in 2020, earlier abundance and tick presence have a negligible effect on *A. americanum* presence in 2024.This assumption can be justified by the fact that the life cycle of *A. americanum* is short, around 2 years only^1^. Finally, we assume that the number of reported deer sightings is proportional to the true abundance of white-tailed deer in the county. That is, we suppose that the reported sightings can be written as a product of the true abundance and an over/under-reporting factor that can potentially depend on the number of humans in the county, the area of the county, the maximum and minimum temperatures, and the total precipitation.

To estimate the expected number of AGS self-reported cases under different levels of white-tailed deer abundance, we used a parametric estimator of the front-door formula based on the plug-in principle and a non-parametric estimation of the joint distribution of exposure and confounders^41^. Specifically, we modeled the distribution of the mediator (*A. americanum* presence, ordered factor with three levels) given the exposure proxy (white-tailed deer reported sightings) and the measured confounders (human population size, county area, maximum and minimum temperature, and total precipitation) using an ordinal logistic regression. The distribution of the outcome variable (AGS self-reported cases) given the mediator, the exposure proxy, and the measured confounders was instead modeled using a negative binomial regression. Details about the regression models and their assessments are reported in the Appendix.

Based on these models, we used the front-door formula to estimate the expected change in number of AGS self-reported cases under five different white-tailed deer abundance scenarios (−50%, +50%, +75%, +100%, +200%). The estimand of interest and the estimators are further described in the Appendix. We obtained 95% confidence intervals using the nonparametric percentile bootstrap with 500 resamples. We then computed the expected difference in the number of AGS self-reported cases in the conterminous United States multiplying these effects by the number of counties.

To test the robustness of our assumptions, we conducted several sensitivity analyses. In particular, we conducted a sensitivity check to assess whether our assumptions of full-mediation and no unmeasured confounding between *A. americanum* presence and AGS self-reported cases were plausible, using data about *I. scapularis.* Furthermore, we re-ran our analyses 1) using the more traditional backdoor criterion, and 2) excluding possible AGS duplicate records. Details about these sensitivity analyses are reported in the Appendix.

All analyses were conducted using R v4.5.1, and RStudio 2025.05.1+513.

## Results

We considered 3,109 US counties or county equivalents, comprising 337,924,709 inhabitants. Overall, we found that 9,481 AGS cases had been self-reported by the end of 2025.

For 1,389 (44.68%) counties there were no records of *A. americanum* presence, for 647 (20.81%) counties the *A. americanum* ticks were reported to be present, while for 1,073 (34.51%) counties the presence of *A. americanum* was established. Counties with presence of *A. americanum* had larger human populations, were of smaller sizes, had higher minimum temperatures, and had more precipitation (see Table 1). The reporting of white-tailed deer sightings was higher in counties where *A. americanum* was present (Table 1). For example, counties with established presence of *A. americanum* had an average of 0.92 white-tailed deer reported sightings per 100 km^2^, versus 0.77 in counties with reported status, and only 0.27 in counties with no records of *A. americanum*. The average number of total AGS self-reported cases per 1,000 inhabitants was also higher in counties with established presence of *A. americanum* (Table 1). In counties with no records of *A. americanum* the average was 0.03 self-reported cases per 1,000 inhabitants, while in counties with reported and established presence of the tick the average was 0.07 and 0.16 per 1,000, respectively.

**Table 1.** Characteristics of 3,109 counties (or county equivalents) in the conterminous United States, by level of *A. americanum* tick presence.

|  | No records<br>(N=1,389) | Reported<br>(N=647) | Established<br>(N=1,073) | Total<br>(N=3,109) |
| --- | --- | --- | --- | --- |
| **Human population (in thousands)** |  |  |  |  |
| Mean (SD) | 89.21 (375.81) | 124.06 (317.67) | 124.64 (301.51) | 108.69 (340.16) |
| Median<br>(Range) | 18.71<br>(0.05, 9,757.18) | 36.55<br>(0.33, 5,009.30) | 32.72<br>(0.63, 5,182.62) | 26.34<br>(0.05, 9,757.18) |
| **Area (km<sup>2</sup>)** |  |  |  |  |
| Mean (SD) | 3,685.97 (4,803.46) | 1,794.40 (1,250.28) | 1,692.22 (902.84) | 2,604.22 (3,443.39) |
| Median<br>(Range) | 2,060.75<br>(7.86, 52,072.73) | 1,512.90<br>(6.54, 9,687.70) | 1,533.63<br>(5.30, 8,742.74) | 1,674.49<br>(5.30, 52,072.73) |
| **Maximum temperature in 2020 (°C)** |  |  |  |  |
| Mean (SD) | 35.60 (3.19) | 35.36 (2.26) | 35.89 (2.08) | 35.65 (2.67) |
| Median<br>(Range) | 35.10<br>(24.38, 47.93) | 35.07<br>(28.10, 44.61) | 35.46<br>(29.11, 45.08) | 35.29<br>(24.38, 47.93) |
| **Minimum temperature in 2020 (°C)** |  |  |  |  |
| Mean (SD) | -16.53 (8.60) | -12.44 (7.75) | -9.16 (5.76) | -13.13 (8.22) |
| Median<br>(Range) | -15.35<br>(-37.30, 5.67) | -11.54<br>(-30.42, 3.85) | -7.92<br>(-26.21, 4.45) | -12.03<br>(-37.30, 5.67) |
| **Total precipitation in 2020 (mm)** |  |  |  |  |
| Mean (SD) | 794.38 (520.52) | 1,190.66 (430.36) | 1,329.37 (358.98) | 1,061.49 (513.89) |
| Median<br>(Range) | 660.82<br>(55.58, 2,941.26) | 1,138.65<br>(187.16, 2,361.11) | 1,379.10<br>(283.75, 2,087.44) | 1,066.02<br>(55.58, 2,941.26) |
| **Number of white-tailed deer reported sightings** |  |  |  |  |
| Mean (SD) | 3.29 (14.38) | 8.56 (25.91) | 9.91 (49.43) | 6.67 (32.93) |
| Median<br>(Range) | 0.00<br>(0.00, 354.00) | 1.00<br>(0.00, 333.00) | 1.00<br>(0.00, 1,324.00) | 1.00<br>(0.00, 1,324.00) |
| **Number of white-tailed deer reported sightings per 100 km<sup>2</sup>** |  |  |  |  |
| Mean (SD) | 0.27 (1.54) | 0.77 (2.87) | 0.92 (5.77) | 0.60 (3.79) |
| Median<br>(Range) | 0.00<br>(0.00, 27.24) | 0.09<br>(0.00, 45.87) | 0.08<br>(0.00, 125.77) | 0.03<br>(0.00, 125.77) |
| **Number of total AGS self-reported cases** |  |  |  |  |
| Mean (SD) | 0.60 (1.97) | 2.58 (5.80) | 6.51 (12.89) | 3.05 (8.54) |
| Median<br>(Range) | 0.00<br>(0.00, 38.00) | 0.00<br>(0.00, 56.00) | 2.00<br>(0.00, 152.00) | 0.00<br>(0.00, 152.00) |
| **Number of total AGS self-reported cases per 1,000 inhabitants** |  |  |  |  |
| Mean (SD) | 0.03 (0.13) | 0.07 (0.20) | 0.16 (0.32) | 0.08 (0.23) |
| Median<br>(Range) | 0.00<br>(0.00, 2.55) | 0.00<br>(0.00, 2.20) | 0.04<br>(0.00, 5.50) | 0.00<br>(0.00, 5.50) |

The association between white-tailed deer reporting, *A. americanum* presence, and AGS self-reported cases is visible when plotting the geographical distribution of these variables. The reporting of white-tailed deer was higher in the South, in the Northeast, and in some areas of the Midwest (Figure 2). White-tailed deer sightings were absent in several areas of the West. Similarly, *A. americanum* ticks were mostly present in the South and in some areas of the Northeast and the Midwest, whereas counties in the West were largely classified as having no records (Figure 3). Counties with the highest rate of self-reported AGS were in the South and in the Midwest, with fewer cases in the West (Figure 4).

**Figure 2.**
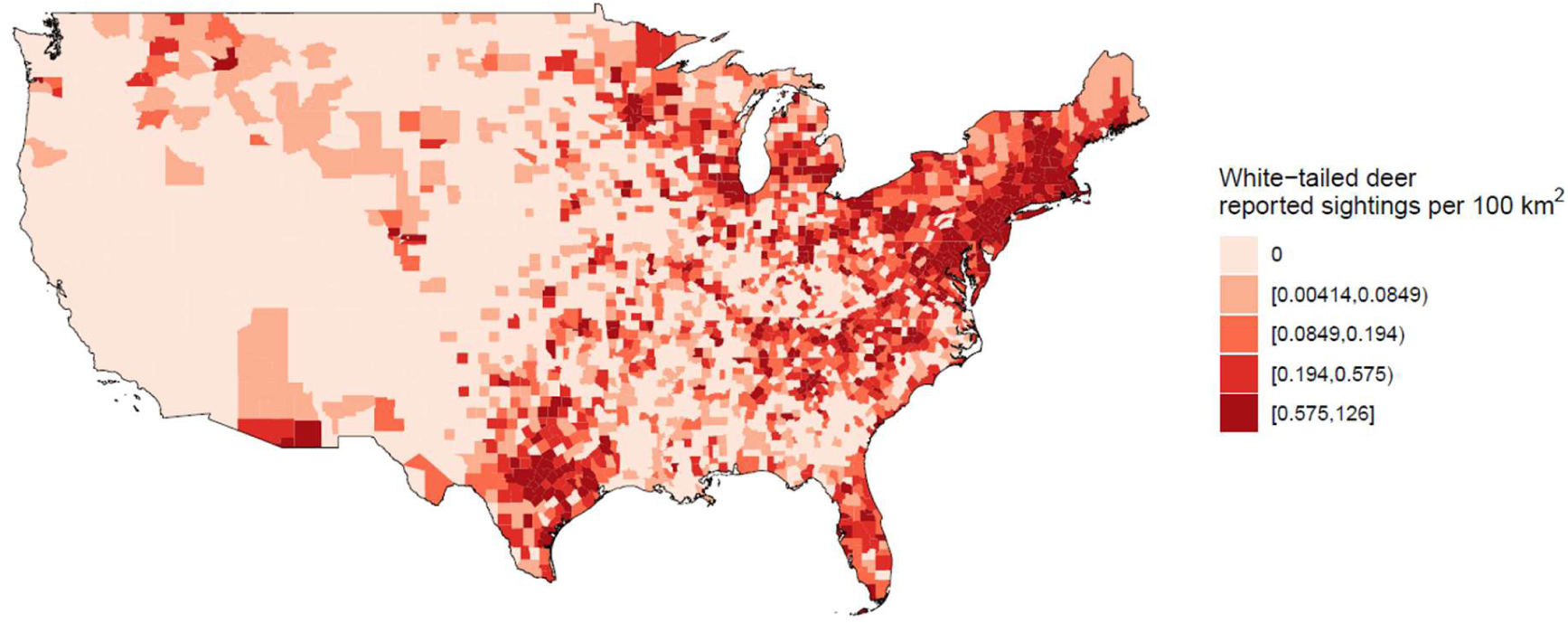
White-tailed deer reported sightings per 100 km^2^ in 2020 by US county (or county equivalent). Data about sightings was obtained from the Global Biodiversity Information Facility (GBIF)^35^, based on records registered through the iNaturalist app^36^.

**Figure 3.**
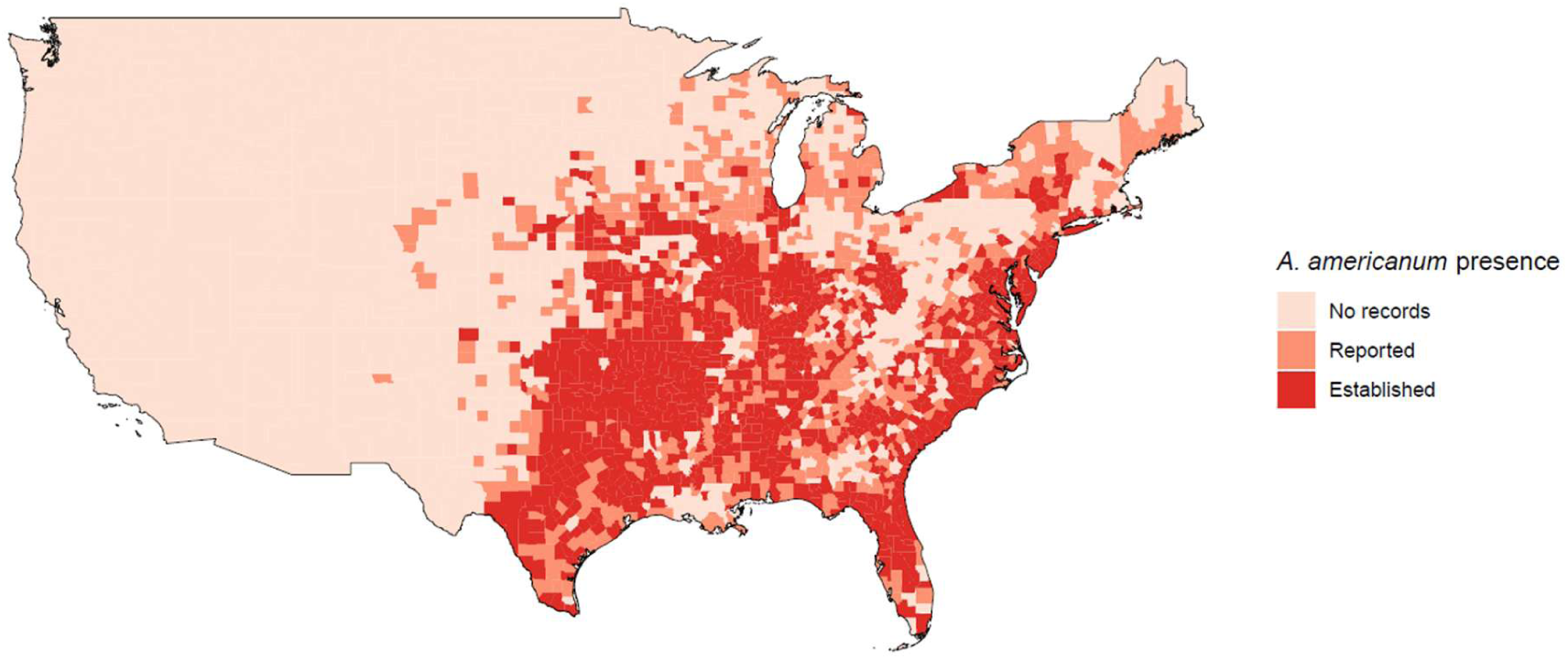
*A. americanum* tick presence by US county (or county equivalent) through 2024. Data were obtained from the CDC surveillance program, which reports the presence of *A. americanum* in three levels: No records, Reported, and Established^33^.

**Figure 4.**
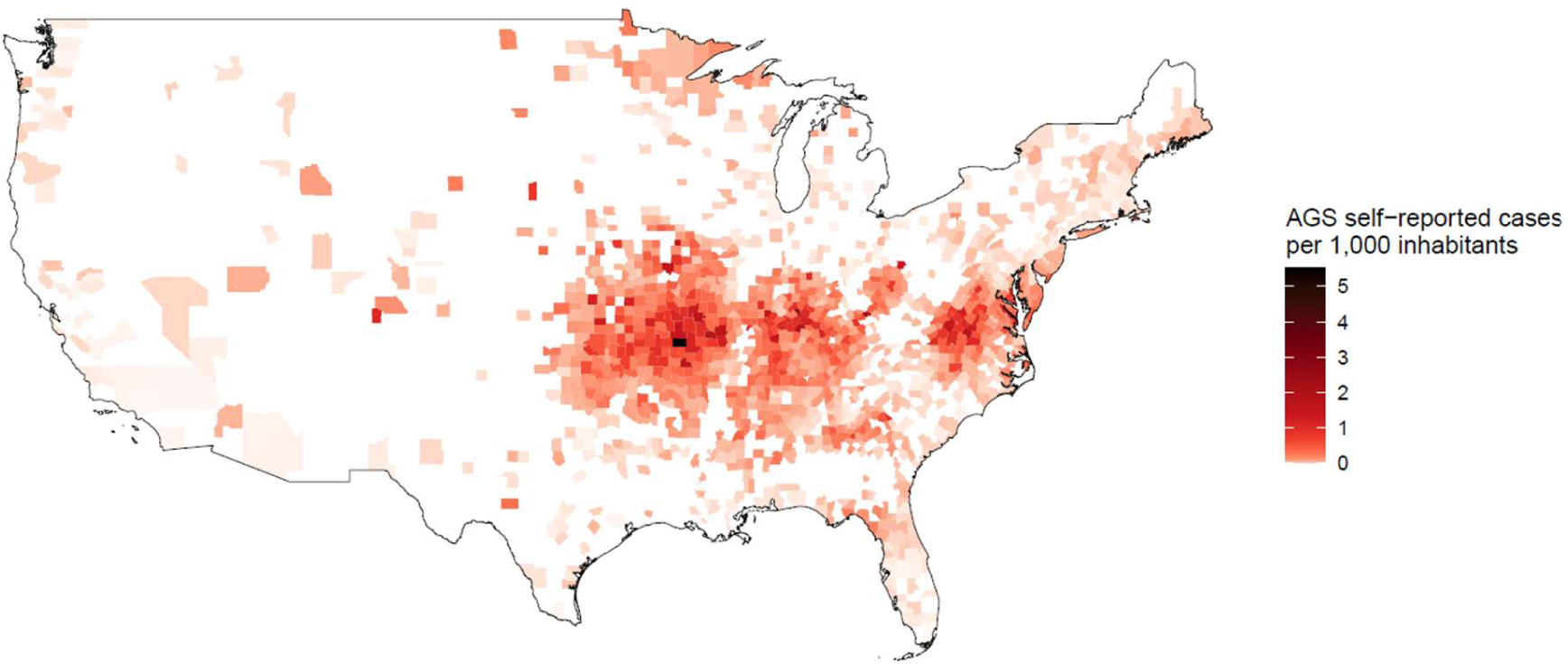
Total alpha-gal syndrome self-reported cases per 1,000 inhabitants by US county (or county equivalent) on 31 December 2025. Data about the number of cases were obtained from a crowdsourced map that collected for years information about self-reported location of individuals with Alpha-gal syndrome^40^.

Results from the regression models are reported in the Appendix. We found that the number of reported white-tailed deer sightings in 2020 was associated with *A. americanum* presence in 2024 (Appendix Table 1). Specifically, we found that the number of white-tailed deer reported sightings was positively associated with the probability of observing *A. americanum*. In turn, higher levels of *A. americanum* presence were associated with a higher number of self-reported AGS cases (Appendix Table 2). Among counties with similar covariate values, those with established *A. americanum* presence were estimated to have approximately twice the expected number of AGS self-reported cases as counties with reported presence, and more than three times the expected number as counties with no records of *A. americanum*.

**Table 2.**
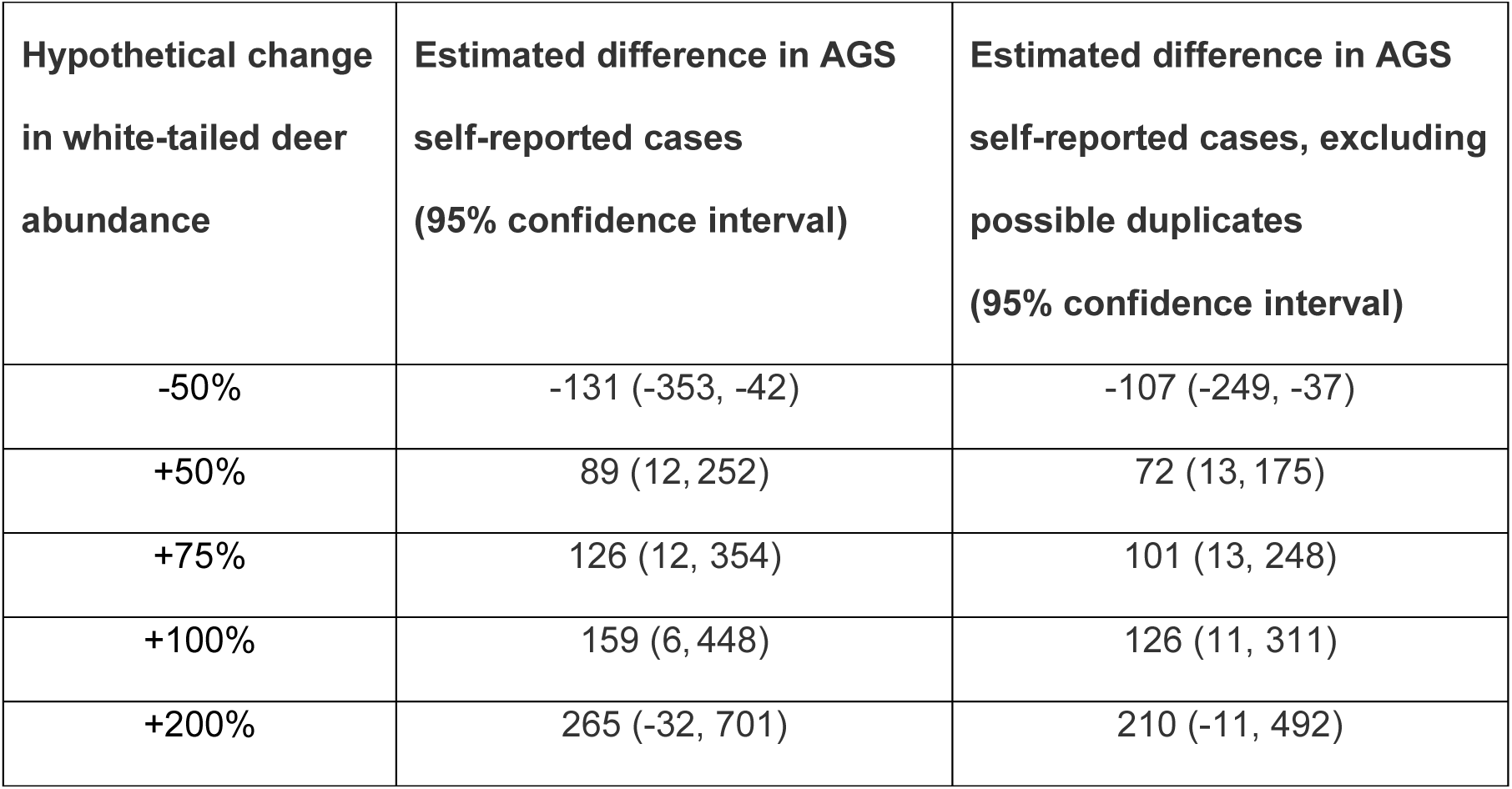
Estimated difference in the number of Alpha-gal syndrome self-reported cases in 2025 in the US under different scenarios of 2020 white-tailed deer abundance. Each scenario corresponds to a change expressed in percentage of the natural white-tailed deer abundance. In the last column are reported results from a sensitivity analysis where possible duplicate AGS records have been excluded. Estimates were obtained using the front-door formula.

| Hypothetical change in white-tailed deer abundance | Estimated difference in AGS self-reported cases (95% confidence interval) | Estimated difference in AGS self-reported cases, excluding possible duplicates (95% confidence interval) |
| --- | --- | --- |
| -50% | -131 (-353, -42) | -107 (-249, -37) |
| +50% | 89 (12, 252) | 72 (13, 175) |
| +75% | 126 (12, 354) | 101 (13, 248) |
| +100% | 159 (6, 448) | 126 (11, 311) |
| +200% | 265 (-32, 701) | 210 (-11, 492) |

Overall, our analysis suggests that increasing white-tailed deer abundance would lead to a higher number of total AGS self-reported cases (Table 2). For example, had the white-tailed deer abundance been halved in 2020, we estimated that there would be 131 (95% CI: 42, 353) fewer AGS self-reported cases in the US in 2025. Conversely, increasing white-tailed deer abundance by 50%, 75%, or 100% would have led to 89 (12, 252), 126 (12, 354) or 159 (6, 448) additional AGS self-reported cases according to our estimates. For the extreme case of a 200% increase in white-tailed deer abundance our estimates had more uncertainty, and the confidence interval included the null value (Table 2). This seems driven by the uncertainty of the regression model in estimating the exposure-mediator relationship in the tail of the exposure distribution, where the association is small and data points are few.

In the sensitivity analysis, excluding possible duplicate records of AGS, we found a total of 8,018 AGS self-reported cases. Overall, this sensitivity analysis gave results similar to the main analysis (Table 2). However, as expected, the estimated effects on AGS self-reported cases were slightly smaller. When using the backdoor criterion, which likely is more prone to bias than the front-door method, estimates had high uncertainty and showed no evidence of an effect (Appendix Table 3). In the sensitivity check, we found no statistically significant difference between counties with established presence of *I. scapularis* and counties where *I. scapularis* was reported (p=0.73), or with counties with no records of *I. scapularis* (p=0.11). This strengthens our belief in the assumption of full mediation and no unmeasured common causes of the variables *A. americanum* presence and AGS self-reported cases.

## Discussion

In this ecological study, we found that a higher number of reported white-tailed deer sightings in 2020 was associated with county-level presence of *A. americanum* in 2024. Furthermore, the county-level presence of *A. americanum* was associated with a higher number of self-reported AGS cases in 2025. These associations are compatible with an effect of white-tailed deer abundance on AGS prevalence in the country.

Our results are consistent with recent theories linking white-tailed deer presence and AGS prevalence^11,20^. They are also consistent with case-control study results suggesting that individuals with AGS were more likely than controls to have lived in areas with deer, such as larger properties, wooded forest, and properties with shrubs and brush^6^. Although in this case-control study, the association between disease and deer sightings had a wide 95% confidence interval, covering the null effect^6^.

Our study has several limitations and our effect estimates should be interpreted cautiously. First, while citizen science efforts provide valuable data that are otherwise difficult to collect, they rely on spontaneous reporting from untrained individuals. Measurement error is almost inevitable. For example, as a proxy for white-tailed deer abundance we used data about reporting from iNaturalist^37^. We assumed that the number of reported deer sightings corresponds to the true abundance of white-tailed deer in the county multiplied by an over/under-reporting factor, which can depend on the number of inhabitants, area of the county, maximum and minimum temperatures, and total precipitation. This assumption likely fails when reporting is affected by other factors as well. Yet, it is reassuring that the geographic distribution derived from iNaturalist reporting data (Figure 1) is quite consistent with information about deer density reported from institutional sources in previous time periods^42^. Also, the outcome variable was the cumulative number of AGS self-reported cases in 2025, obtained from an online map^40^. No specific instructions about the location required were provided to the users, and these numbers might suffer from inaccuracies in the data entering process. We conducted a sensitivity analysis to mitigate the impact of errors in the data entering process. Yet, it is possible that our outcome variable does not directly reflect a quantity of public health interest, for example due to underreporting. We emphasize, however, that the geographic distribution from the self-reported data (Figure 3) closely resembles the official data about suspected cases from the CDC for the period 2017-2022^15^, suggesting a close relationship between actual number of cases and self-reported cases.

Second, in our main analysis based on the front-door criterion, we assumed that *A. americanum* presence is the full mediator of the effect of white-tailed deer abundance on AGS self-reported cases. We also required that there are no unmeasured common causes of this mediator and the exposure/outcome. The plausibility of these assumptions is complicated by the lack of time granularity for the *A. americanum* presence variable. To reduce concerns about violation of our assumptions, we adjusted our analysis for important drivers of *A. americanum* presence (i.e., extreme temperatures, area of the county, total precipitation). We also conducted a sensitivity check using data on another tick species. This sensitivity check found no evidence of violation for the full mediation assumption and was compatible with no unmeasured common causes between the full mediator and the outcome. Despite these reassuring results, our analysis might still be biased. For example, tick presence was only available as a categorical variable, and might fail to capture all effects of white-tailed deer abundance on AGS self-reported cases.

Finally, we relied on common regression models, which invoke parametric restrictions, and assumed that counties were independent observations. Because some modelling choices were data-driven, our estimates should be considered as exploratory, and statistical inference should be interpreted with caution.

AGS is a growing public health concern in the United States^15^. Recently, a fatal anaphylactic reaction related to AGS was reported^20^. AGS has received wide media coverage^43^ and has also led to controversial debates in bioethics^44^. Despite this, epidemiological data about AGS are scarce^15^. Our work gives preliminary evidence that an increase in the number of white-tailed deer can lead to a higher number of individuals developing AGS. Further studies are needed to better quantify the effect of white-tailed deer abundance on AGS prevalence and plan appropriate public health interventions. Answering this research question is particularly important now, as the number of white-tailed deer is increasing, especially towards the north^11,17,20^, due to human modification of the landscape, hunting regulation, reduction of predators, and climate change^45,46^.

## Data Availability

The data supporting the findings of this study are publicly available. Estimates for the number of persons living in each US county in 2024 according to the U.S. Census Bureau were downloaded from StatsAmerica' s Download Center29. TIGER/Line shapefiles and information about the total area of each county from the U.S. Census Bureau were obtained using the "tigris" R package30. Data about temperature and precipitation for each county were obtained from the National Oceanic and Atmospheric Administration using the "EpiNOAA" R package31. The estimated presence of A. americanum through 2024 in each US county was downloaded from the Lone Star Tick Surveillance website of the Centers for Disease Control and Prevention (CDC)33. Data on the number of reported human observations of white-tailed deer were downloaded from the Global Biodiversity Information Facility (GBIF)36. Data about the number of self-reported AGS cases were obtained from a crowdsourced map40.

## Acknowledgements

During the preparation of this manuscript the authors used ChatGPT to assist with formatting and minor language editing. The authors reviewed and edited the content as needed and take full responsibility for the content of the publication.

## Authorship contribution statement

MP and MJS conceptualized and designed the study. MP collected the data, with support from LC. MP performed the analyses. MP and LC wrote the first draft. All authors revised the manuscript critically for important intellectual content, approved the final version, and agreed to be accountable for all aspects of the work.

## Data availability statement

The data supporting the findings of this study are publicly available. Estimates for the number of persons living in each US county in 2024 according to the U.S. Census Bureau were downloaded from StatsAmerica’ s Download Center^29^. TIGER/Line shapefiles and information about the total area of each county from the U.S. Census Bureau were obtained using the “tigris” R package^30^. Data about temperature and precipitation for each county were obtained from the National Oceanic and Atmospheric Administration using the “EpiNOAA” R package^31^. The estimated presence of *A. americanum* through 2024 in each US county was downloaded from the Lone Star Tick Surveillance website of the Centers for Disease Control and Prevention (CDC)^33^. Data on the number of reported human observations of white-tailed deer were downloaded from the Global Biodiversity Information Facility (GBIF)^36^. Data about the number of self-reported AGS cases were obtained from a crowdsourced map^40^.

## References

1. Shishido AA, Wormser GP. A review of alpha-gal syndrome for the infectious diseases practitioner. Open Forum Infect Dis. 2025;12:ofaf430.

2. Kepley CL, Wang Y, Yelton A, Siebert ER, Iweala OI. Ticked off: Allergic effector cells in the pathogenesis of alpha-gal syndrome. Curr Allergy Asthma Rep. 2025;25:57.

3. Commins SP. Diagnosis & management of alpha-gal syndrome: lessons from 2,500 patients. Expert Rev Clin Immunol. 2020;16:667–77.

4. Platts-Mills TAE, Commins SP, Biedermann T, van Hage M, Levin M, Beck LA, et al. On the cause and consequences of IgE to galactose-α-1,3-galactose: A report from the National Institute of Allergy and Infectious Diseases Workshop on Understanding IgE-Mediated Mammalian Meat Allergy. J Allergy Clin Immunol. 2020;145:1061–71.

5. Young I, Prematunge C, Pussegoda K, Corrin T, Waddell L. Tick exposures and alpha-gal syndrome: A systematic review of the evidence. Ticks Tick Borne Dis. 2021;12:101674.

6. Kersh GJ, Salzer J, Jones ES, Binder AM, Armstrong PA, Choudhary SK, et al. Tick bite as a risk factor for alpha-gal-specific immunoglobulin E antibodies and development of alpha-gal syndrome. Ann Allergy Asthma Immunol. 2023;130:472–8.

7. Carson AS, Gardner A, Iweala OI. Where’s the beef? Understanding allergic responses to red meat in alpha-gal syndrome. J Immunol. 2022;208:267–77.

8. Binder AM, Commins SP, Altrich ML, Wachs T, Biggerstaff BJ, Beard CB, et al. Diagnostic testing for galactose-alpha-1,3-galactose, United States, 2010 to 2018. Ann Allergy Asthma Immunol. 2021;126:411–416.e1.

9. Choudhary S, Iweala OI, Addison CT, Commins SP. Tick salivary extract induces alpha-gal allergy in alpha-gal deficient mice. J Allergy Clin Immunol. 2019;143:AB252.

10. Naseem Z, Muhammad A, Chatterjee A, Rubio-Tapia A. Alpha-gal syndrome: Recognizing and managing a tick-bite-related meat allergy. Cleve Clin J Med. 2025;92:311–9.

11. Platts-Mills TAE, Gangwar RS, Workman L, Wilson JM. The immunology of alpha-gal syndrome: History, tick bites, IgE, and delayed anaphylaxis to mammalian meat. Immunol Rev. 2025;332:e70035.

12. Chung CH, Mirakhur B, Chan E, Le QT, Berlin J, Morse M, et al. Cetuximab-induced anaphylaxis and IgE specific for galactose-alpha-1,3-galactose. N Engl J Med. 2008;358:1109–17.

13. Commins SP, Satinover SM, Hosen J, Mozena J, Borish L, Lewis BD, et al. Delayed anaphylaxis, angioedema, or urticaria after consumption of red meat in patients with IgE antibodies specific for galactose-alpha-1,3-galactose. J Allergy Clin Immunol. 2009;123:426–33.

14. Van Nunen SA, O’Connor KS, Clarke LR, Boyle RX, Fernando SL. An association between tick bite reactions and red meat allergy in humans. Med J Aust. 2009;190:510– 1.

15. Thompson JM, Carpenter A, Kersh GJ, Wachs T, Commins SP, Salzer JS. Geographic distribution of suspected alpha-gal syndrome cases - United States, January 2017-December 2022. MMWR Morb Mortal Wkly Rep. 2023;72:815–20.

16. Commins SP, Karim S. Development of a novel murine model of alpha-gal meat allergy. J Allergy Clin Immunol. 2017;139:AB193.

17. Paddock CD, Yabsley MJ. Ecological havoc, the rise of white-tailed deer, and the emergence of Amblyomma americanum-associated zoonoses in the United States. Curr Top Microbiol Immunol. 2007;315:289–324.

18. Bloemer SR, Zimmerman RH, Fairbanks K. Abundance, attachment sites, and density estimators of lone star ticks (Acari: Ixodidae) infesting white-tailed deer. J Med Entomol. 1988;25:295–300.

19. Mount GA, Haile DG, Barnard DR, Daniels E. New version of LSTSIM for computer simulation of Amblyomma americanum (Acari: Ixodidae) population dynamics. J Med Entomol. 1993;30:843–57.

20. Platts-Mills TAE, Workman LJ, Richards NE, Wilson JM, McFeely EM. Implications of a fatal anaphylactic reaction occurring 4 hours after eating beef in a young man with IgE antibodies to galactose-α-1,3-galactose. J Allergy Clin Immunol Pract. 2025;13:3422–4.

21. Hernán MA, Robins JM. Causal Inference: What if. Boca Raton: Chapman & Hall/CRC; 2020. 352 p.

22. Pearl J. Causality: Models, Reasoning and Inference. 2nd ed. Cambridge University Press; 2009. 464 p.

23. Pearl J, Mackenzie D. The Book of Why: The New Science of Cause and Effect. Penguin UK; 2018. 432 p.

24. Pearl J. Mediating Instrumental Variables. Technical Report R-210, Cognitive Systems Laboratory, UCLA Computer Science Department [Internet]. 1993; Available from: Available at: https://ftp.cs.ucla.edu/pub/stat_ser/r210.pdf

25. Inoue K, Ritz B, Arah OA. Causal effect of chronic pain on mortality through opioid prescriptions: Application of the front-door formula. Epidemiology. 2022;33:572–80.

26. Piccininni M, Kurth T, Audebert HJ, Rohmann JL. The effect of mobile stroke unit care on functional outcomes: An application of the front-door formula. Epidemiology. 2023;34:712–20.

27. Saunders EF, Sohail H, Myles DJ, Charnetzky D, Ayres BN, Nicholson WL, et al. Alpha-gal syndrome after Ixodes scapularis Tick Bite and statewide surveillance, Maine, USA, 2014-2023. Emerg Infect Dis. 2025;31:809–13.

28. Butler WK, Oltean HN, Dykstra EA, Saunders E, Salzer JS, Commins SP. Onset of alpha-gal syndrome after Tick Bite, Washington, USA. Emerg Infect Dis. 2025;31:829– 32.

29. Download center [Internet]. 2025 [cited 2025 Dec 14]. Available from: https://www.statsamerica.org/downloads/default.aspx

30. Walker K. Tigris: An R package to access and work with geographic data from the US census bureau. R J. 2016;8:231.

31. Applied Research Center Documentation. EpiNOAA overview [Internet]. 2026 [cited 2026 June 22]. Available from: https://arc.ncics.org/r-package-overview/

32. Kessler WH, Ganser C, Glass GE. Modeling the distribution of medically important tick species in Florida. Insects. 2019;10:E190.

33. CDC. Lone Star tick surveillance [Internet]. Ticks. 2025 [cited 2025 Dec 14]. Available from: https://www.cdc.gov/ticks/data-research/facts-stats/lone-star-tick-surveillance.html

34. Census Bureau. Change to county-equivalents in the State of Connecticut [Internet]. Federal Register. 2022. Available from: https://www.federalregister.gov/d/2022-12063

35. GBIF: The Global Biodiversity Information Facility (2025). What is GBIF? [Internet]. 2025 [cited 2025 Dec 14]. Available from: https://www.gbif.org/what-is-gbif

36. GBIF.org (31 December 2025). GBIF Occurrence Download [Internet]. 2025. Available from: 10.15468/dl.5nnept

37. About iNaturalist [Internet]. iNaturalist. 2025 [cited 2025 Dec 14]. Available from: https://www.inaturalist.org/pages/about

38. Alpha-gal Information. About Us [Internet]. 2022 [cited 2025 Dec 14]. Available from: https://alphagalinformation.org/about-us/

39. Alpha-gal Information. Where Does AGS Occur? [Internet]. 2025 [cited 2025 Dec 14]. Available from: https://alphagalinformation.org/where/

40. Where in the world is Alpha Gal? [Internet]. 2025 [cited 2025 Dec 31]. Available from: https://www.zeemaps.com/map?group=555038

41. Fulcher IR, Shpitser I, Marealle S, Tchetgen Tchetgen EJ. Robust inference on population indirect causal effects: the generalized front door criterion. J R Stat Soc Series B Stat Methodol. 2020;82:199–214.

42. Hanberry B, Hanberry P. Rapid digitization to reclaim thematic maps of white-tailed deer density from 1982 and 2003 in the conterminous US. PeerJ. 2020;8:e8262.

43. Wells P. Why Is Martha’s Vineyard Going Vegan? It’s All About Tick Bites. The New York Times [Internet]. 2025 [cited 2025 Dec 28]; Available from: https://www.nytimes.com/2025/08/12/dining/marthas-vineyard-alpha-gal-tick-bites.html

44. Crutchfield P, Hereth B. Beneficial bloodsucking. Bioethics. 2025;39:772–81.

45. The Wildlife Society. NE Section Position Statement on Managing Chronically Overabundant Deer [Internet]. 2018 [cited 2026 July 22]. Available from: https://wildlife.org/ne-section-position-statement-on-managing-chronically-overabundant-deer/

46. Dawe KL, Boutin S. Climate change is the primary driver of white-tailed deer (Odocoileus virginianus) range expansion at the northern extent of its range; land use is secondary. Ecol Evol. 2016;6:6435–51.

